# The research fatigue and beneficence scale: development and validation in a nationwide cohort of transgender women in the United States and Puerto Rico

**DOI:** 10.64898/2026.04.13.26350829

**Authors:** Meg Stevenson, Sari L. Reisner, Ceza Pontes, Sabriya Linton, Annick Borquez, Asa E. Radix, Jason S. Schneider, The ENCORE Study Group, Erin Cooney, Andrea L. Wirtz

## Abstract

Transgender women are routinely recruited for HIV prevention research and describe feeling over-researched, undervalued, and disconnected from the benefits of research. Research fatigue refers to the adverse impacts of research participation from the volume, frequency, or intensity of research engagement. Research beneficence, an underdeveloped construct, refers to perceptions that research participation is empowering, appreciated, and beneficial to individuals and communities. This study sought to develop and psychometrically evaluate a research fatigue and beneficence scale and examine associations with cohort retention and study procedures among transgender women in the US and Puerto Rico.

We developed a novel 7-item measure of research fatigue and beneficence informed by prior literature and qualitative work with transgender women. We assessed internal consistency reliability, factor structure, convergent and divergent validity, and predictive validity with 6-month study retention outcomes and procedures among 2189 transgender women enrolled in a US nationwide cohort (April 2023–December 2024) for the full 7-item research fatigue and beneficence scale, a 4-item research beneficence subscale, and a single-item research fatigue measure.

Research beneficence items demonstrated good internal consistency (0.78) and excellent model fit. Research fatigue and beneficence varied by race/ethnicity with participants of color reporting both greater empowerment and greater concerns about community-level benefits. The item “I feel that I am asked to participate in research too frequently” was associated with lower 6-month retention, greater survey missingness, and preference for less invasive HIV testing modalities.

Findings highlight multiple dimensions of research experience and the need for reduced participant burden, culturally tailored study designs, and intentional dissemination efforts to improve participant-centered research practices.

## Introduction

With overdue attention to minimizing the burden of research participation on study participants, the concept of “research fatigue” among frequently researched populations has gained attention across multiple fields.^1-7^ For the present study, we define research fatigue as the adverse impacts of research participation on study participants due to its frequency or intensity. These adverse impacts often include feeling under-valued or over-researched; believing that the research does not benefit participants or their communities; and becoming unwilling, reluctant, or disengaged as a result of prior research experiences.^1-3^

We define research beneficence, the conceptual antonym to research fatigue, as participants feeling empowered, appreciated, and benefitted through their involvement in research. The concept of research beneficence is rooted in the Belmont report’s fundamental ethical principle that research should benefit participants and society to the maximum extent possible.^8^ In this paper, we extend this ethical research principle by considering research participants’ perception of beneficence and exploring how investigators might mitigate research fatigue by enhancing both direct and indirect benefits to participants and their communities.

Understanding research fatigue and beneficence from a human subjects protection perspective and biomedical ethics standpoint is necessary for designing studies that appropriately address participant protections, compensation, and dissemination of findings. These findings are also important from a scientific perspective, as high levels of research fatigue may impact recruitment, retention, data quality, and induce selection bias. Likewise, understanding participant perceptions of research beneficence may strengthen both human subjects’ protections and study rigor. Some populations, including small communities (e.g. gender diverse communities, small indigenous or ethnic groups), those exposed to specific risk factors (e.g. sex work, homelessness, drug use, migration), experiencing chronic illnesses (or associated occupational exposures), or those living in areas prone to natural disasters, industrial contamination, or other structural vulnerabilities, are at higher risk of research fatigue. As research increasingly occurs through online and digital modalities, allowing broader access to small or historically underrepresented populations, the potential for research fatigue is likely to grow, making addressing these issues more important than ever.^4^

Recent literature on research fatigue has provided guidelines or best practices to reduce its impact on study participants.^2-5,9^ Recommended strategies include prioritizing dissemination of results and focusing on outcomes that then communities of interest feel are valuable to them.^9^ Researchers increasingly report difficulty recruiting participants from highly researched communities, and that their participation is sometimes reluctant or disengaged. These researchers report that communities perceive their involvement is fruitless despite the frequency of participation, and relay second-hand accounts from community members that indicate that they feel over researched and that despite being frequently involved in research, they don’t see their participation as translating to changes that benefit them; that “people are always researching [them] but nothing changes.” These themes are notably consistent across diverse populations—including young people living with HIV, Rwandan refugees, energy-impacted communities, Indigenous peoples, transgender women, and people who use illicit substances^1,3,5-7^—highlighting the broad relevance of research fatigue. A smaller body of qualitative research documents these experiences from the perspectives of highly researched populations themselves. Individuals describe feeling like “guinea pigs” or “lab rats,” perceiving little resulting benefit for their communities, lacking clarity about how research findings are used, and feeling jaded by the sheer volume of studies.^10,11^

Transgender women are one such highly-researched population, yet only recently have investigators begun to research transgender women separately from men who have sex with men.^12,13^ Concurrently, transgender women in the US experience increasing disenfranchisement in healthcare and other aspects of society.^14^ These factors highlight not only the research burden on transgender women, but the conditions which prime them to experience research fatigue and the importance of prioritizing research beneficence.

Qualitative research among transgender women echoes the same sentiments expressed by other highly-researched populations. In a qualitative study among transgender women in Lima, Peru, exploring research mistrust in PrEP research,^11^ 86 transgender women described an increasing burden of research participation alongside limited improvements in HIV outcomes; dismissal from researchers and medical professionals about the medical and social realities of taking PrEP; and persistent barriers to PrEP access despite ongoing involvement in clinical trials.^11,15^ Qualitative work with transgender women in the US echoes these findings. In a study led by members of the present study team interviewing participants who had previously engaged in HIV research, participants critiqued lengthy and repetitive surveys, expressed mistrust regarding the lack of transparency about how their data would be used, and feeling “used” or like a “guinea pig.”^15^ An additional qualitative study on trans erasure in healthcare, participants reported that despite participating in health research, their research contributions are often not incorporated into textbooks or healthcare protocols, or are conflated with the experiences of gay men, contributing to feelings of alienation and stigmatization.^16^

In response to these longstanding concerns, a 2018 article published recommendations for ethical approaches to recruitment strategies and collaboration with transgender participants in academic research, citing these documented frustrations, heightened community alienation, and history of objectification and delegitimization of transgender participants by researchers.^17^ These recommendations emphasized an awareness and appreciation of trans history; transparency about study outcomes, goals, and potential benefits to research participants; and careful attention to language used. Building upon this foundation, the current study sought to develop and psychometrically evaluate a quantitative measure of research fatigue and beneficence among transgender women, a population highly burdened by the HIV epidemic and frequently engaged in HIV research; and to examine associations with cohort retention and study procedures among transgender women in the US. The goal is to systematically assess these constructs and provide a validated tool that can help researchers to identify, reduce, and proactively address research fatigue while enhancing beneficence in future studies.

## Methods

### Study Design

This analysis was conducted using baseline data from a nationwide epidemiologic cohort of 2,504 transgender women and trans feminine individuals in the US, including Puerto Rico. The primary aims of the parent study are to estimate HIV incidence, identify predictors of HIV acquisition, and assess engagement in HIV prevention services. Detailed study methods have been published previously.^18^ Briefly, study participants were enrolled between April 2023 – December 2024 and completed visits every 6 months. This was a hybrid cohort: participants could enroll and participate across the US and Puerto Rico through remote, digital methods, while participants located near study sites in Atlanta, Baltimore, Durham, Miami, New York City, and San Diego had the option to enroll and/or participate in-person at study hubs. Each semi-annual assessment consisted of a self- or interviewer-administered biobehavioral survey and HIV test. To maximize response to HIV testing, participants had the options to provide (1) an oral fluid specimen for laboratory-based antibody testing; (2) dried blood spot (DBS) for laboratory-based antibody/antigen testing, (3) staff-administered rapid test performed at the study hub, or (4) submit laboratory results from HIV testing performed for clinical purposes in the last 30 days. Participants received $15 for a completed biobehavioral survey at each timepoint and $65 for a completed HIV test at each timepoint, administered through their choice of digital gift cards or physical reloadable debit card.

### Study Sample

Individuals were eligible if they resided in any of the 50 US states, Puerto Rico, or Washington DC; were not living with HIV (biomarker-confirmed); could communicate in English and/or Spanish; were ages 18 years or older; and self-reported a current transgender women, woman, female, or transfeminine identity, and were assigned male sex at birth.

### Measures

#### Research Fatigue & Beneficence Scale

Seven items were developed to assess for research fatigue and beneficence (RFB). Item development drew upon a literature review; previous qualitative work by the authors with this study population exploring factors influencing engagement in HIV research and healthcare;^19^ informal discussions with participants in previous studies;^20^ and consultation with transgender research team and advisory board members. The 7-item measure was included on the baseline survey for all cohort participants and is displayed in Table 2. Three items assessed research fatigue (items 3, 5, and 6), and 4 measured research beneficence (items 1, 2, 4, and 7). Responses were assessed using a 5-point Likert scale ranging from strongly disagree to strongly agree, along with the option “prefer not to answer.” Research fatigue items (3, 5, and 6) were reverse coded and all seven items were summed together to create a research fatigue and beneficence score. The four research beneficence items were summed separately to create a research beneficence score. The three research fatigue items were summed separately to create a research fatigue score. All three scales were assessed for goodness of fit using a Cronbach’s alpha of .70 as acceptable and a Cronbach’s alpha of .80 as good.^21^ Individual RFB items were also considered for assessment as single-factor predictors of study outcomes.

#### Correlates

Demographics examined for relationship with the RFB items included race, ethnicity, previous participation in research led by the study team, lifetime and past-year history of HIV testing, lifetime homelessness, lifetime history of STI testing, online vs. in person study participation, food security, geographic location, educational attainment, employment status, income, gender identity, sexual orientation, unstable housing, immigration status, internet access, material hardship, incarceration history, self-reported health, healthcare experiences, health insurance status, PTSD, HIV prevention history, lifetime sex work, PrEP use, substance use,^22,23^ gender-affirming care history, gender euphoria,^24^ legal name/gender marker change, Adverse Childhood Experiences (ACEs),^25^ Positive Childhood Experiences (PCEs),^26^ experiences of bullying, vicarious trauma, dignity,^27^ and flourishing.^28^ Relationships were determined using chi-square tests to assess for statistically significant differences in proportions.

We created a variable of survey missingness for follow up study assessments, in which the numerator represents the number of missing responses in the survey and the denominator represents all survey items which were shown to all participants regardless of skip patterns.

#### Variables for Convergent and Divergent Validity

Variables for convergent and divergent validity were selected based on their conceptual relationship to research fatigue and beneficence and previous literature and assessed using Pearson’s *r* correlation coefficients. Validated measures and their Cronbach’s alphas among our sample were: Transgender Pride (α=0.70), Social Support (α=0.80), Community Connectedness (α=0.67), Anticipated Discrimination (α=0.84), and Psychological Distress via the Kessler-6,^29^ (α=0.85).

#### Variables for Predictive Validity

Selected HIV testing modality (DBS or oral fluid) was assessed at baseline and 6-month follow-up. We examined completion of study visit components at the 6-month follow up visit, including the 6-month survey and HIV test completion. Survey missingness was assessed as the number of missing responses in the survey (numerator) and all survey items which were shown to all participants regardless of skip patterns (denominator).

### Analytic sample

The baseline sample of 2,504 participants was restricted to participants with 1 or fewer missing items on the research fatigue and beneficence measures. Participants who selected “prefer not to answer” to more than 1 scale item were excluded from this analysis (n=28). Participants who completed their baseline survey prior to May 30^th^ 2023 (n=287) were also excluded from this analysis due to a programming error preventing all response options from being displayed. Our final analytic sample for this analysis was 2,189 participants.

### Statistical approach

Univariate statistics were computed to assess the frequency and distribution of all variables included in the analysis. Of the three conceptualized scales, scales with an acceptable or good Cronbach’s alpha were summarized with means and standard deviations, and confirmatory factor analyses (CFA) were conducted.

#### Internal consistency reliability

Cronbach’s alpha was assessed for both the full research fatigue and beneficence scale, and separately for the research beneficence and research fatigue subscales. For assessing internal validity of the scale and subscales via Cronbach’s alpha, a cut-off of 0.70 was used as acceptable and 0.80 as good.^21^ We assessed for differences in experiences of research fatigue and beneficence by race/ethnicity through chi-square tests conducted for individual items, research fatigue & beneficence 7-item scale score, and research beneficence 4-item scale score.

#### Factor analysis

Confirmatory factor analysis was performed and with good model fit indices defined as Comparative Fit Index (CFI) over 0.95, Tucker-Lewis Index (TLI) over 0.90, Standardized Root Mean Square Residual (SRMR) over 0.08, and with Root Mean Squared Error of Approximation (RMSEA) defined as excellent at 0.01, good at 0.05, and acceptable at 0.08.^30^ All statistical analyses were performed using Stata version 18. Statistical significance was determined using a p-value of 0.05.

#### Convergent and divergent validity

Convergent and divergent validity with other validated psychometric scales were also assessed (Transgender Pride, Social Support, Community Connectedness, Anticipated Discrimination, and Psychological Distress via Kessler-6).^29,31,32^ Descriptive tables presenting the distribution of responses have been collapsed into 3-point Likert scales for ease of interpretation, but the full 5-point Likert scales were retained for validation analyses.

#### Predictive Validity

Predictive validity was assessed through longitudinal analyses to determine associations between baseline research fatigue and beneficence measures and completion of study visit components at the 6 month follow up visit and 6-month survey missingness. Because we used this missingness item to determine the validity of participants at the baseline encounter for enrollment in the cohort, we only use this item as an outcome at 6-month follow up. HIV testing and survey completion were defined as binary variables in which participants did or did not fully complete their survey and HIV tests and follow up timepoints – again because these components were required for enrollment into the cohort, they could not be used as an outcome at the baseline encounter. HIV testing modality was used as an outcome at both baseline and 6-month follow up timepoints, measuring the predictive value of the single-item research fatigue measure on the proportion of participants who opted out of antigen/antibody testing via DBS in favor of antibody testing via oral fluid.

### Ethical considerations

The Johns Hopkins Single Institutional Review Board reviewed and approved this study (IRB00355445) and served as the institutional review board of record for all partner institutions in this multisite study. As a minimal risk study, participants provided consent in English or Spanish using an oral consent form in web-based format prior to initiating research activities.

## Results

Table 1 displays demographic characteristics of the analytic sample. The sample was 64% monoracial white participants; 48% had a lifetime experience of homelessness and 57% had low or very low food security.

**Table 1:** Demographic characteristics of a nationwide cohort of transgender women in the US and Puerto Rico (N=2189)

|  | *n* | % |
| --- | --- | --- |
| **Age (mean, IQR) (N=2189)** | 34.3 | 25 - 40 |
| **Race and Ethnicity (N=2189)** |  |  |
| Non-Hispanic White | 1393 | 64% |
| Non-Hispanic Black | 152 | 7% |
| Hispanic White | 139 | 6% |
| Hispanic Black | 24 | 1% |
| Non-Hispanic Multiracial or Other | 280 | 13% |
| Hispanic Multiracial or Other | 201 | 9% |
| **Education (n=2171)** |  |  |
| High school diploma/GED or less | 484 | 22% |
| Some college or higher | 1687 | 77% |
| **Employment Status (n=2146)** |  |  |
| Unemployed | 865 | 40% |
| Part-time | 362 | 17% |
| Full-time | 919 | 43% |
| **Region (n=2188)** |  |  |
| Mid-Atlantic | 335 | 15.3% |
| Midwest | 452 | 20.7% |
| New England | 97 | 4.4% |
| Puerto Rico | 10 | 0.5% |
| South | 558 | 25.5% |
| Southwest | 202 | 9.2% |
| West | 534 | 24.4% |
| **Lifetime homelessness (n=2163)** |  |  |
| No | 1130 | 52.2% |
| Yes | 1033 | 47.8% |
| **Food Security (n=2091)** |  |  |
| High or marginal | 902 | 43.1% |
| Low or very low | 1189 | 56.9% |

**Table 2:**
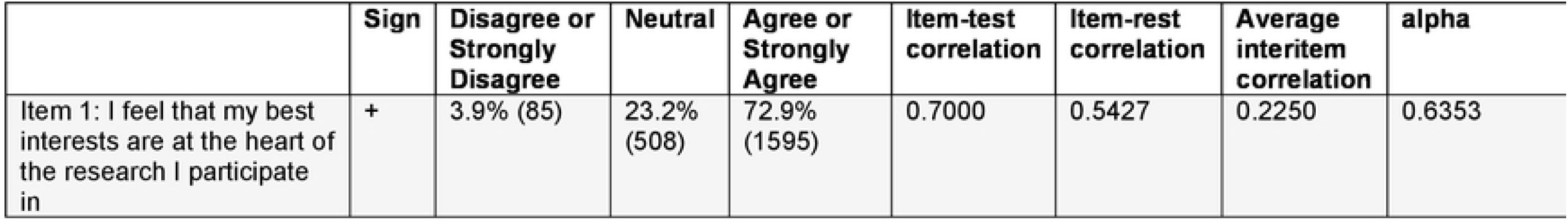
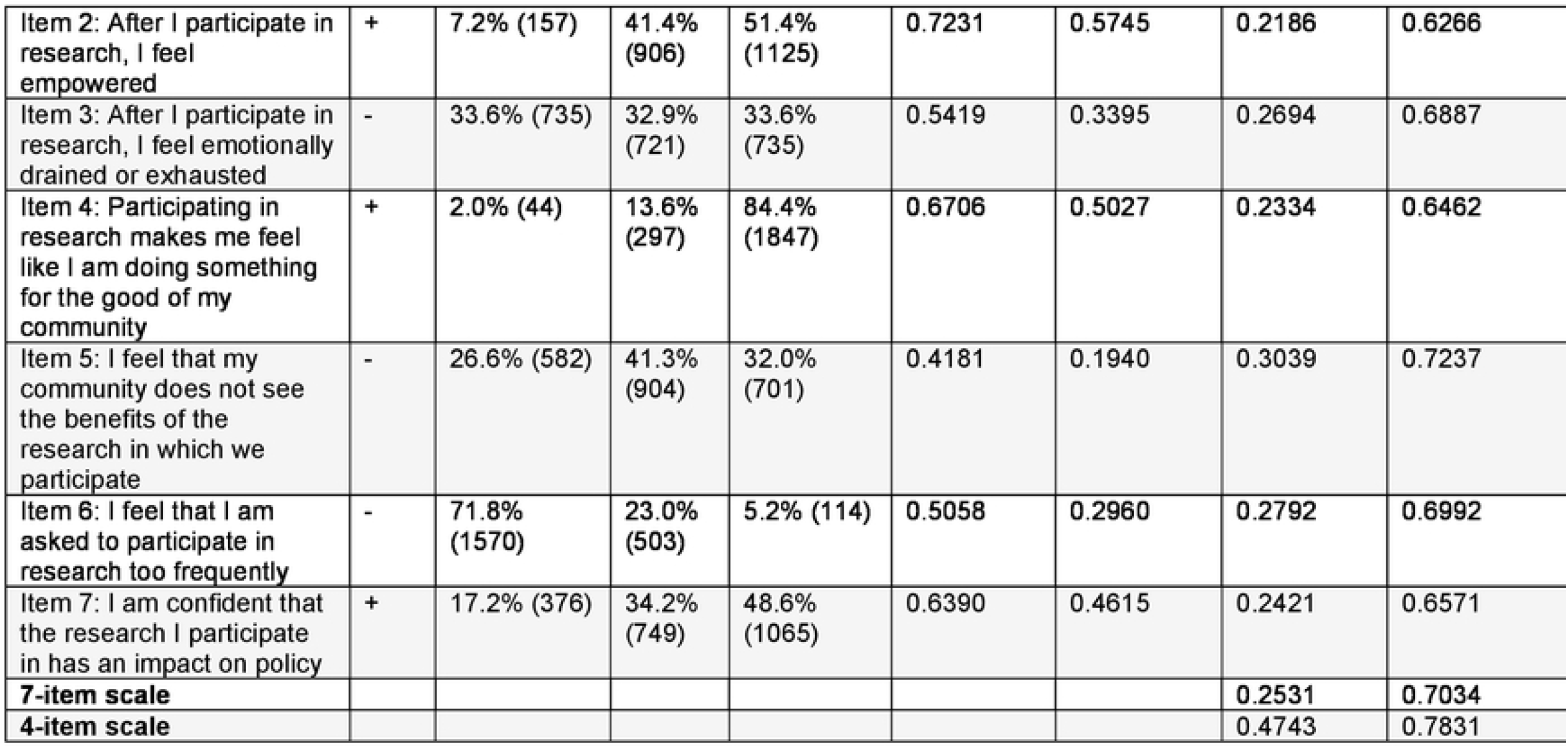
Research fatigue and beneficence scale item distribution and internal consistency among a nationwide cohort of lransgender women in the US and Puerto Rico (N=2189)

**Table 3:** Model fit statistics of two scales measuring research beneficence among a nationwide cohort of transgender women in the US and Puerto Rico (N=2189)

|  | CFI | TLI | RMSEA | SRMR |
| --- | --- | --- | --- | --- |
| 7-Item | 0.974 | 0.951 | 0.063 | 0.035 |
| 4-Item | 0.999 | 0.993 | 0.036 | 0.007 |

Table 2 displays the seven items in the research fatigue and beneficence scale, the response distributions in the study sample, and internal consistency results. Endorsement of individual research fatigue survey items varied from 5.2% (Item 6: I feel that I am asked to participate in research too frequently) to 33.6% (Item 3: After I participate in research, I feel emotionally drained or exhausted). Endorsement of individual research beneficence survey items varied from 48.6% (Item 7: I am confident that the research I participate in has an impact on policy) to 84.4% (Item 4: Participating in research makes me feel like I am doing something for the good of my community).

The full scale demonstrated good internal consistency (α=0.70) when all items were included.^21,33^ A 4-item subscale comprised of the research beneficence items (items 1, 2, 4, and 7) demonstrated stronger internal consistency (α=0.78). A 3-item subscale including the research fatigue items (items 3, 5, and 6) had poor internal consistency (α=0.47) [data not displayed], and thus was not further assessed.

Fit statistics for the proposed 7- and 4-item scales are displayed, which indicate excellent model fit. Despite otherwise robust model fit statistics, in the full 7-item scale, item 6 (“I feel I am asked to participate in research too frequently” ) had no unique variance apart from the other items, so results should be interpreted with caution. A 6 item scale removing item 6 resulted in comparable internal consistency with the full scale (α=0.69).

Table 4 shows Pearson *r* correlations for both the 4- and 7-item scales with other validated instruments included in our survey. Both scales are positively associated with transgender pride, social support, and community connectedness, and inversely associated with anticipated discrimination and psychological distress.

**Table 4:** Convergent and divergent validity: Pearson *r* correlations of research fatigue and beneficence scales with conceptually-selected constructs among a nationwide cohort of transgender women in the US and Puerto Rico (N=2189)

|  | 4-Item Research Beneficence | 7-Item Research Fatigue & Beneficence | Transgender Pride | Social Support | Community Connectedness | Anticipated Discrimination | Psychological Distress |
| --- | --- | --- | --- | --- | --- | --- | --- |
| 4-Item Research Beneficence | 1.000 |  |  |  |  |  |  |
| 7-Item Research Fatigue & Beneficence | 0.859*** | 1.000 |  |  |  |  |  |
| Transgender Pride | 0.223*** | 0.165*** | 1.000 |  |  |  |  |
| Social Support | 0.079** | 0.104*** | 0.083** | 1.000 |  |  |  |
| Community Connectedness | 0.138*** | 0.171*** | 0.284*** | 0.225*** | 1.000 |  |  |
| Anticipated Discrimination | -0.065* | -0.121*** | -0.055* | -0.183*** | -0.069* | 1.000 |  |
| Psychological Distress (K-6) | -0.079** | -0.101*** | -0.105*** | -0.210*** | -0.149*** | 0.333*** | 1.000 |

\*p<0.05 \*\*p<.01 \*\*\*p<0.001

Table 5 displays experiences of research fatigue and beneficence by race/ethnicity and age. Items 2 (After I participate in research, I feel empowered), 5 (I feel that my community does not see the benefits of the research in which we participate), 6 (I feel that I am asked to participate in research too frequently), and 7 (I am confident that the research I participate in has an impact on policy) differed significantly by race. Items 1 (I feel that my best interests are at the heart of the research I participate in) and 5 (I feel that my community does not see the benefits of the research in which we participate) differed significantly by age.

**Table 5:** Research fatigue and beneficence measures by race/ethnicity and age group among a nationwide cohort of transgender women in the US and Puerto Rico (N=2189)

| | Non-Hispanic White (n=1393) | Person of Color (n=796) | *p-value* ($\chi^2$) | Age 18-24 (n=X) | Age 25+ (n=X) | *p-value* ($\chi^2$) |
| --- | --- | --- | --- | --- | --- | --- |
| **I feel that my best interests are at the heart of the research I participate in** |  |  | 0.523 |  |  | 0.001 |
| (Strongly) Disagree | 53 (3.8%) | 32 (4.0%) |  | 7 (1.6%) | 78 (4.5%) |  |
| Neutral | 334 (24.0%) | 174 (21.9%) |  | 90 (20%) | 418 (24.1%) |  |
| (Strongly Agree) | 1391 (72.2%) | 589 (74.1%) |  | 354 (78.5%) | 1239 (71.4%) |  |
| **After I participate in research, I feel empowered** |  |  | 0.001 |  |  | 0.303 |
| (Strongly) Disagree | 103 (7.4%) | 54 (6.8%) |  | 36 (8.0%) | 121 (7.0%) |  |
| Neutral | 614 (44.1%) | 292 (36.6%) |  | 173 (38.4%) | 733 (42.3%) |  |
| (Strongly) Agree | 674 (48.5%) | 449 (56.6%) |  | 242 (53.7%) | 881 (50.8%) |  |
| **After I participate in research, I feel emotionally drained or exhausted** |  |  | 0.574 |  |  | 0.439 |
| (Strongly) Disagree | 456 (32.7%) | 278 (34.9%) |  | 163 (36.1%) | 571 (32.9%) |  |
| Neutral | 463 (33.2%) | 258 (32.4%) |  | 144 (31.9%) | 577 (33.2%) |  |
| (Strongly) Agree | 474 (34.0%) | 260 (32.7%) |  | 145 (32.1%) | 589 (33.9%) |  |
| **Participating in research makes me feel like I am doing something for the good of my community** |  |  | 0.572 |  |  | 0.919 |
| (Strongly) Disagree | 31 (2.2%) | 13 (1.7%) |  | 8 (1.8%) | 36 (2.1%) |  |
| Neutral | 185 (13.3%) | 112 (14.1%) |  | 61 (13.5%) | 236 (13.6%) |  |
| (Strongly) Agree | 1175 (84.5%) | 670 (84.3%) |  | 382 (84.7%) | 1463 (84.3%) |  |
| **I feel that my community does not see the benefits of the research in which we participate** |  |  | <0.001 |  |  | 0.032 |
| (Strongly) Disagree | 404 (29.0%) | 178 (22.4%) |  | 129 (28.7%) | 453 (26.1%) |  |
| Neutral | 612 (44.0%) | 291 (36.7%) |  | 200 (44.4%) | 703 (40.5%) |  |
| (Strongly) Agree | 375 (27.0%) | 325 (41.0%) |  | 121 (26.9%) | 579 (33.4%) |  |
| **I feel that I am asked to participate in research too frequently** |  |  | <0.001 |  |  | 0.103 |
| (Strongly) Disagree | 1048 (75.2%) | 521 (65.8%) |  | 342 (75.8%) | 1227 (70.8%) |  |
| Neutral | 303 (21.8%) | 200 (25.2%) |  | 89 (19.7%) | 414 (23.9%) |  |
| (Strongly) Agree | 42 (3.0%) | 71 (9.1%) |  | 20 (4.4%) | 93 (5.4%) |  |
| **I am confident that the research I participate in has an impact on policy** |  |  | <0.001 |  |  | 0.111 |
| (Strongly) Disagree | 289 (20.8%) | 87 (10.9%) |  | 92 (20.4%) | 284 (16.4%) |  |
| Neutral | 516 (37.0%) | 233 (29.3%) |  | 154 (34.1%) | 595 (34.3%) |  |
| (Strongly) Agree | 588 (42.2%) | 475 (59.8%) |  | 206 (45.6%) | 857 (49.4%) |  |
| **Mean (SD) Scale Scores** |  |  |  |  |  |  |
| 7 item | 5.8 (3.8) | 6.0 (3.8) |  | 6.2 (3.5) | 5.8 (3.9) |  |
| 4 item | 10.9 (2.6) | 11.4 (2.8) |  | 11.1 (2.6) | 11.0 (2.7) |  |

**Table 6:** Single-item research fatigue measure associations with study retention activities in a nationwide cohort of transgender women in the US and Puerto Rico (N=2189)

|  |  |  | I feel that I am asked to participate in research too frequently |  |  |  |  |  |  |
| --- | --- | --- | --- | --- | --- | --- | --- | --- | --- |
| *Characteristic* | Overall sample: | | (Strongly) Disagree | | Neutral | | (Strongly) Agree | | p-value ($\chi^2$) |
|  | n | % | n | % | n | % | n | % |  |
| **Testing Modality (Baseline)** |  |  |  |  |  |  |  |  | **0.001** |
| DBS | 1766 | 84% | 1283 | 84% | 411 | 84% | 70 | 70% |  |
| Oral Fluid | 347 | 16% | 237 | 16% | 78 | 16% | 30 | 30% |  |
| **Testing Modality (6-Month)** |  |  |  |  |  |  |  |  | **0.008** |
| DBS | 1492 | 78% | 1099 | 79% | 303 | 78% | 51 | 65% |  |
| Oral Fluid | 410 | 22% | 286 | 21% | 96 | 23% | 28 | 35% |  |
| **Survey Missingness (6-Month)** |  |  |  |  |  |  |  |  | **0.010** |
| mean % responses missing |  | 1.7% |  | 1.4% |  | 2.0% |  | 4.1% |  |
| **Survey Completion (6-Month)** |  |  |  |  |  |  |  |  | **<0.001** |
| % completion | 1981 | 91% | 1438 | 92% | 452 | 90% | 91 | 81% |  |
| **HIV Testing Completion (6-Month)** |  |  |  |  |  |  |  |  | **0.002** |
| % completion | 1693 | 77% | 1226 | 78% | 395 | 79% | 72 | 64% |  |

Because item 6, *I feel that I am asked to participate in research too frequently*, had no unique variance apart from the other 7-item scale items, yet has a strong conceptual relationship with the outcomes of interest conceptually and prioritized by research participants in the literature, we assessed item 6 separately as a single-item predictor of study participation and attrition at the 6-month assessment. Those who endorsed item 6 were more likely to select less-invasive antibody HIV testing completed with oral fluid specimens (reference: antigen-antibody HIV testing completed via dried blood spot) at both baseline and 6-month timepoints; less likely to be retained in the study at 6 months follow up; and had higher average missingness at their 6-month follow up survey.

## Discussion

In this analysis of a novel measure of research fatigue and beneficence among a nationwide cohort of transgender women in the US, our full 7-item research beneficence and fatigue scale and 4-item research beneficence scale had good psychometric properties. The 3-item scale restricted to research fatigue items had poor internal consistency, thus we do not suggest it for use measuring experiences of research fatigue. We propose both the 7-item research fatigue and beneficence scale and the 4-item research beneficence scale for further exploration as appropriate tools to characterize experiences related to research among highly-researched populations.

We found most participants did not feel that they were asked to participate in research too frequently. However, experiences of research fatigue were common, including feeling that their participation did not benefit them and their communities or influence policy. Participants who reported being asked to participate in research too frequently were less likely to be retained at follow up. Concurrently, participants in our sample reported high levels of research beneficence.

Experiences of research fatigue and beneficence varied by ethnoracial identity. Transgender women of color were less likely to report seeing benefits from the research in which they participate and more likely to feel over-approached for research. However, participants of color also reported higher levels of empowerment due to their research participation. These findings highlight the multidimensionality of research participation experiences. Individual participants may simultaneously feel empowered and exhausted, or, as in the case of transgender women of color in this sample, perceive that research impacts policy but does not benefit their own communities. Interpreted in the context of existing literature, this finding may reflect a belief that research disproportionately benefits other members of the broader LGBTQ+ community, such as gay men or White transgender women, rather than transgender women of color.^12,16,17^ It is also important to consider the intersectional experiences of transgender women of color, as people of color have long experienced mistrust of and mistreatment by the research and medical communities.^34^

Experiences of research fatigue and beneficence also varied by age group, with older participants less likely to report a perception that their best interests were at the heart of the research in which they participate. They were also more likely to report that their community does not see the benefits of the research in which they participate. These findings suggest a need to prioritize older transgender people in research and design studies exploring their unique needs.

The association of being asked to participate in research too frequently with study attrition has important implications for scientific inference, particularly if these experiences are disproportionately experienced by marginalized groups or groups with highest burden of health outcomes of interest. As online research continues to expand and reach historically underrepresented populations, planning for retention during the study design stage should explicitly consider research fatigue. Participants who felt over-approached were more likely to choose antibody HIV testing via oral fluid specimens, which may be perceived as less invasive than collection of antibody/antigen dried-blood spots, but may not detect very recent infections and is not preferred for those most vulnerable to HIV acquisition.^35^ Reducing the research burden on the participant so those who would most benefit from the most sensitive testing methods are not too overburdened to complete them is critical. Conversely, offering multiple, less invasive biological testing options may help reduce overall research fatigue and associated attrition among highly researched populations. We managed this by offering both antibody testing via oral fluid and antigen/antibody testing via DBS, and counseling participants that the offered DBS testing is better at detecting recent HIV acquisitions and allowing the participants to select the testing methodology which felt most appropriate for their perceived risk and tolerance for blood-based testing. These findings may apply to other biological specimen collection methods and should be considered for other types of research. Similarly, participants who reported being asked to participate in research too frequently had increased survey missingness at follow up, emphasizing the need to carefully consider time burden and potential fatigue associated with lengthy or sensitive measures when designing surveys.

Our findings also highlight the importance of disseminating research results to participants. Consistent with prior literature, feeling that participation in research has a real-world impact on one’s community and the broader policy and scientific environment is a crucial component of mitigating research fatigue and increasing feelings of research beneficence. The vast majority of this study sample felt that participating in research made them feel that they were doing something for the good of their community, yet a much smaller proportion endorsed the statement “I am confident that the research I participate in has an impact on policy” and even fewer disagreed with the statement “I feel that my community does not see the benefits of the research in which we participate.” While participants felt that the research being conducted was of value to them personally and to their wider communities, they also felt disconnected from the impacts of the research. Increasing transparency and communication about study impacts and sharing research results may help mitigate fatigue and enhance beneficence. Similarly, creating and meaningfully engaging community advisory boards in research and continuing these partnerships through community action should be considered a core component of research with highly-researched populations.

Our study has several limitations. First, this study was conducted among transgender women without HIV who are currently engaged in research, and their experiences with research may not be reflective of the broader population in the United States. Secondly, due to the self-reflective nature of a research study evaluating research fatigue, the potential impacts of social desirability bias must be considered. Despite assurance that survey responses will not influence one’s participation in the study, it is possible that participants who valued the financial incentive or HIV testing services would be less likely to report negative attitudes towards research. Finally, this validation study requires replication; we suggest exploring the application of the full scale in other contexts and among other populations.

To our knowledge, this manuscript is among the first to report on the development of quantitative measures to assess research fatigue and beneficence in any population; if these scales are adopted by other research teams, this will allow for comparative analyses between groups as opposed to the purely qualitative literature which exists currently. This manuscript also expands the discussion beyond negative experiences of research fatigue to include positive experiences of research beneficence, which we hope will support improved research practices for these populations moving forward. Further research is needed evaluate the performance of this measure in other populations, as well as among transgender women in different research contexts.

## Conclusions

Transgender women have received increasing attention in research as the health inequities experienced by these communities have received more attention and support. Our findings contribute to evidence that research engaging transgender women can feel opaque, exploitative, or fruitless, and that these perceptions may contribute to study attrition. By examining how these experiences differ by race and by validating two versions of a scale assessing research beneficence, with or without items on research fatigue, we offer new tools for understanding and improving participant-centered research. We recommend these scales be delivered to and tested with diverse populations and settings to evaluate their broader utility.

## Data Availability

Data is available upon reasonable request to the corresponding author.

## Supplemental

**Table S1:** Four-item scale measuring research fatigue and beneficence among a nationwide cohort of transgender women in the US and Puerto Rico (N=2189)

|  | Observations | Sign | Item-test correlation | Item-rest correlation | Average inter-item correlation | alpha |
| --- | --- | --- | --- | --- | --- | --- |
| Item 1: I feel that my best interests are at the heart of the research I participate in | 2186 | + | 0.7853 | 0.5994 | 0.4671 | 0.7245 |
| Item 2: After I participate in research, I feel empowered | 2186 | + | 0.8230 | 0.6619 | 0.4274 | 0.6913 |
| Item 4: Participating in research makes me feel like I am doing something for the good of my community | 2186 | + | 0.7748 | 0.5823 | 0.4787 | 0.7337 |
| Item 7: I am confident that the research I participate in has an impact on policy | 2188 | + | 0.7310 | 0.5140 | 0.5242 | 0.7677 |
| Test Scale |  |  |  |  | 0.4743 | 0.7831 |

**Table S2:**
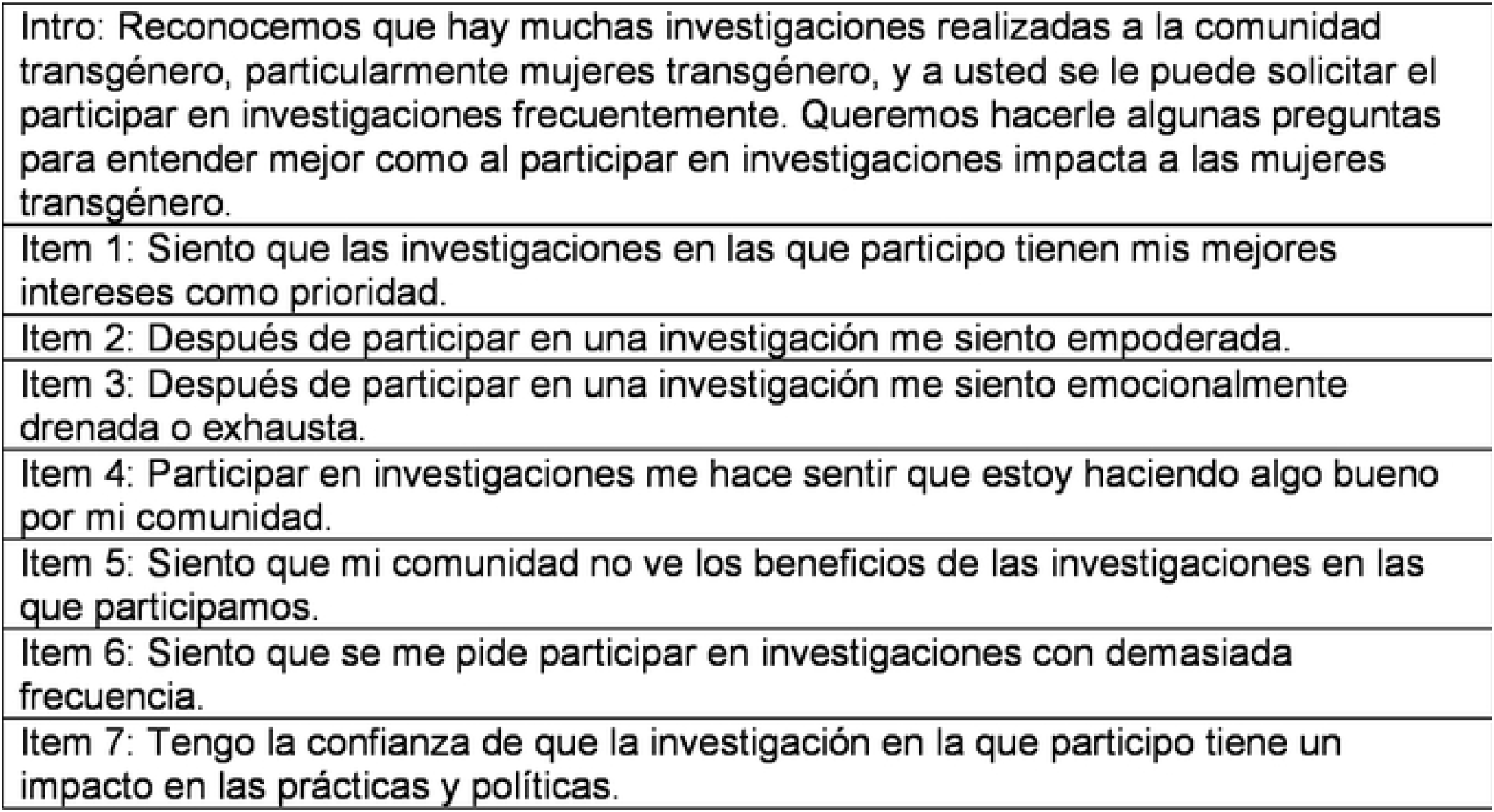
Spanish language research fatigue and beneficence scale.

## Notes

### Competing Interest Statement

TCP is a consultant for ViiV Healthcare and received an honorarium for a lecture provided to Merck & Co staff. ALW, TCP, MS, CP, and EEC receive research funding to their institutions from ViiV Healthcare. SR receives royalties from McGraw Hill for editing textbook on transgender health and is a Member of Board of Directors for WPATH.

### Clinical Protocols

https://www.researchprotocols.org/2024/1/e59846

### Funding Statement

Yes

